# Multimodal MALDI imaging mass spectrometry for improved diagnosis of melanoma

**DOI:** 10.1101/2022.11.29.22282868

**Authors:** Wanqiu Zhang, Nathan Heath Patterson, Nico Verbeeck, Jessica L. Moore, Alice Ly, Richard M. Caprioli, Bart De Moor, Jeremy L. Norris, Marc Claesen

**Affiliations:** ESAT-STADIUS, KU Leuven, Leuven, Belgium; Aspect Analytics NV, C-mine 12, 3600 Genk, Belgium; Frontier Diagnostics, LLC, Nashville, Tennessee, USA; Mass Spectrometry Research Center, Department of Biochemistry, Vanderbilt University, Nashville, Tennessee, USA

## Abstract

Imaging mass spectrometry (IMS) provides promising avenues to augment histopathological investigation with rich spatio-molecular information. We have previously developed a classification model to differentiate melanoma from nevi lesions based on IMS protein data, a task that is challenging solely by histopathologic evaluation.

Most IMS-focused studies collect microscopy in tandem with IMS data, but this microscopy data is generally omitted in downstream data analysis. Microscopy, nevertheless, forms the basis for traditional histopathology and thus contains invaluable morphological information. In this work, we developed a multimodal classification pipeline that uses deep learning, in the form of a pre-trained artificial neural network, to extract the meaningful morphological features from histopathological images, and combine it with the IMS data.

To test whether this deep learning-based classification strategy can improve on our previous results in classification of melanocytic neoplasia, we utilized MALDI IMS data with collected serial H&E stained sections for 331 patients, and compared this multimodal classification pipeline to classifiers using either exclusively microscopy or IMS data. The multimodal pipeline achieved the best performance, with ROC-AUCs of 0.968 vs. 0.938 vs. 0.931 for the multimodal, unimodal microscopy and unimodal IMS pipelines respectively. Due to the use of a pre-trained network to perform the morphological feature extraction, this pipeline does not require any training on large amounts of microscopy data. As such, this framework can be readily applied to improve classification performance in other experimental settings where microscopy data is acquired in tandem with IMS experiments.

## Introduction

Worldwide, skin cancer is the most common and the most deadly cancer in the white-skinned population.^1,2^ Currently, the diagnosis of melanoma is most frequently reliant upon histopathologic evaluation by dermatopathologists, which is inherently challenging. Several studies have reported significant discordance among experts in the diagnosis of challenging melanocytic lesions.^3,4^ For some challenging lesions, even experienced dermatopathologists had difficulty precisely or reproducibly classifying them into a certain category. Therefore, melanocytic lesions are the most common type of lesions that require a second opinion in pathology. ^5^ However, second opinions can be problematic, and often do not yield the needed accuracy.

One strategy to assist with melanoma diagnosis is by incorporating molecular technologies, to move to a more objective, evidence-based assessment. Imaging Mass Spectrometry (IMS) has been employed increasingly in clinical research and integrated into molecular pathology to complement routine histopathology evaluation. ^6^ The success of IMS is owed to its ability to characterize tissue microenvironments and provide a crucial link between tissue morphology and its molecular physiology. Many studies using IMS on the skin have already shown the potential in diagnosis of melanoma. ^7–15^

In a previous study,^8^ we utilized matrix-assisted laser desorption ionization imaging mass spectrometry (MALDI IMS) technology to classify melanoma and nevi lesions, with the results showing high concordance (97.6% sensitivity and 96.4% specificity) with triconcordant histopathological evaluation by board-certified dermatopathologists. MALDI IMS is a beneficial alternative technology for skin cancer diagnosis as it uses tissue sections prepared in a similar manner as standard histopathology with minimal sample material for *in situ* analysis (a single 6 µm tissue section) and can a provide spatial information when using a serial section for histopathological annotation.^8^

In our previous study, the pipeline we constructed solely used MALDI IMS data to predict the sample classification. However, for each of the IMS experiments, the pathologists used an H&E stain of the neighboring tissue section to guide the histopathological annotations. These H&E images provide crucial morphological information that pathologists use to diagnose melanoma lesions. By targeting the analytical measurement to regions with the highest probability of malignancy, we strengthen the classification algorithm. We hypothesize that a classification model that combines the H&E imaging data, which is the basis of histopathological diagnosis, with MALDI IMS data, an objective but highly accurate means of melanoma classification, that the overall classification accuracy could be further improved. Multimodal imaging is a powerful tool which integrates information from different imaging techniques, and the resulting integrated datasets may open new opportunities for data mining. This can be beneficial for MALDI IMS data, as they contain a substantial amount of chemical information but not enough readily available morphological knowledge from the tissue.^16^ A multimodal MALDI IMS strategy often involves careful data acquisition, registration and computational data analysis to get the most/best of the different modalities.^16^ In this way, complementarities and correlations from both modalities can be explored, providing better evidence towards biological understanding.

Currently, there are very few studies combining both spatial and chemical modalities, i.e., histopathological data and IMS data, to improve the downstream classification task (i.e., diagnosis). At present, there is one study that reports the synergy of metabolomics data from MALDI IMS and morphometry from histopathological data to improve the classification of kidney cancer.^17^ This study was performed on a large patient cohort (n = 853) using FFPE tissue samples; when the classifier was trained on combined or multimodal datasets (morphometric and metabolite), it outperformed the classifiers trained on the individual data for each tumor subtype. This study shows the potential of using both histopathology and IMS data to improve the downstream diagnosis and tumor subtyping.

In comparison to the work by Prade et al. which used a computer-assisted morphometry image analysis algorithm,^18^ we applied a pre-trained deep learning-based model to extract the morphological features from the histopathology dataset in this current study. Specifically, we adapted the pre-trained contrastive self-supervised learning model SimCLR,^19^ pre-trained on 57 histopathology datasets without any labels, ^20^ to extract the high-level morphological features of the histopathological images previously only used to guide the MALDI IMS data acquisition.^8^

In the machine learning field, the number of labeled data is found to have a positive correlation with task performance. However, the most tedious and time-consuming step is to annotate the histological data using medical image analysis applications. As labeled data is not often available, many approaches have been proposed to mitigate this bottleneck. For example, unsupervised learning^21^ and its subclass self-supervised learning do not require annotations of the data, whereas semi-supervised learning uses partially labeled data. Interestingly, self-supervised approaches obtain ‘labels’ from the data itself and predict part of the data from other parts. Instead of focusing on finding specific patterns from the data like unsupervised learning often does, e.g., via clustering, self-supervised learning methods concentrate on recovering whole or part of some features from its original input, while still fitting in the type of supervised settings.^22^

In this paper we adopted the pre-trained contrastive self-supervised learning model from Ciga et al. and applied it on our previously collected 331 patients histopathology datasets.^8^ Whereas in Al-Rohil et al., the histopathological images were used to only guide MALDI-IMS data acquisition, here we also used them in the multimodal data analysis to improve the downstream classification task. Our method successfully extracts relevant and highly qualitative morphological features from the histological images, without requiring any training nor labels. Finally, we combine the extracted morphological feature with MALDI IMS data in the downstream classification task to distinguish melanoma from nevus. We compare the classification performance of the unimodal and multimodal strategies. Our multimodal approach shows improvement over the unimodal MALDI IMS classification results, as reported in our previous work.^8^ Our approach shows the potential of the synergism of spatial proteomics and morphology in (melanoma) cancer diagnosis.

## Materials and Methods

### Tissue Collection

The tissue samples used in this study were the same cohort as used in Al-Rohil et al.. Deidentified FFPE skin biopsies from patients diagnosed with invasive melanoma and nevi were provided by the co-operating academic institutions and private practices. All samples were collected between 2010 and 2016.

The review boards of Indiana University (IRB 1910325060), Vanderbilt University (IRB 030220), Duke University (IRB Pro00102363) and Sterling Institutional Review Board (Atlanta, GA) determined this study exempt from IRB review under the terms of the US Department of Health and Human Services Policy for Protection of Human Research Subjects at 45 CFR §46.104(d).

### Histopathology Sample Preparation and Data Acquisition

Serial sections (6 µm) were cut from deidentified patient biopsy FFPE tissue blocks and used for histopathology and mass spectrometry analysis. Figure 1(a) illustrates the workflow of histology-guided IMS sample preparation and processing.

**Figure 1:**
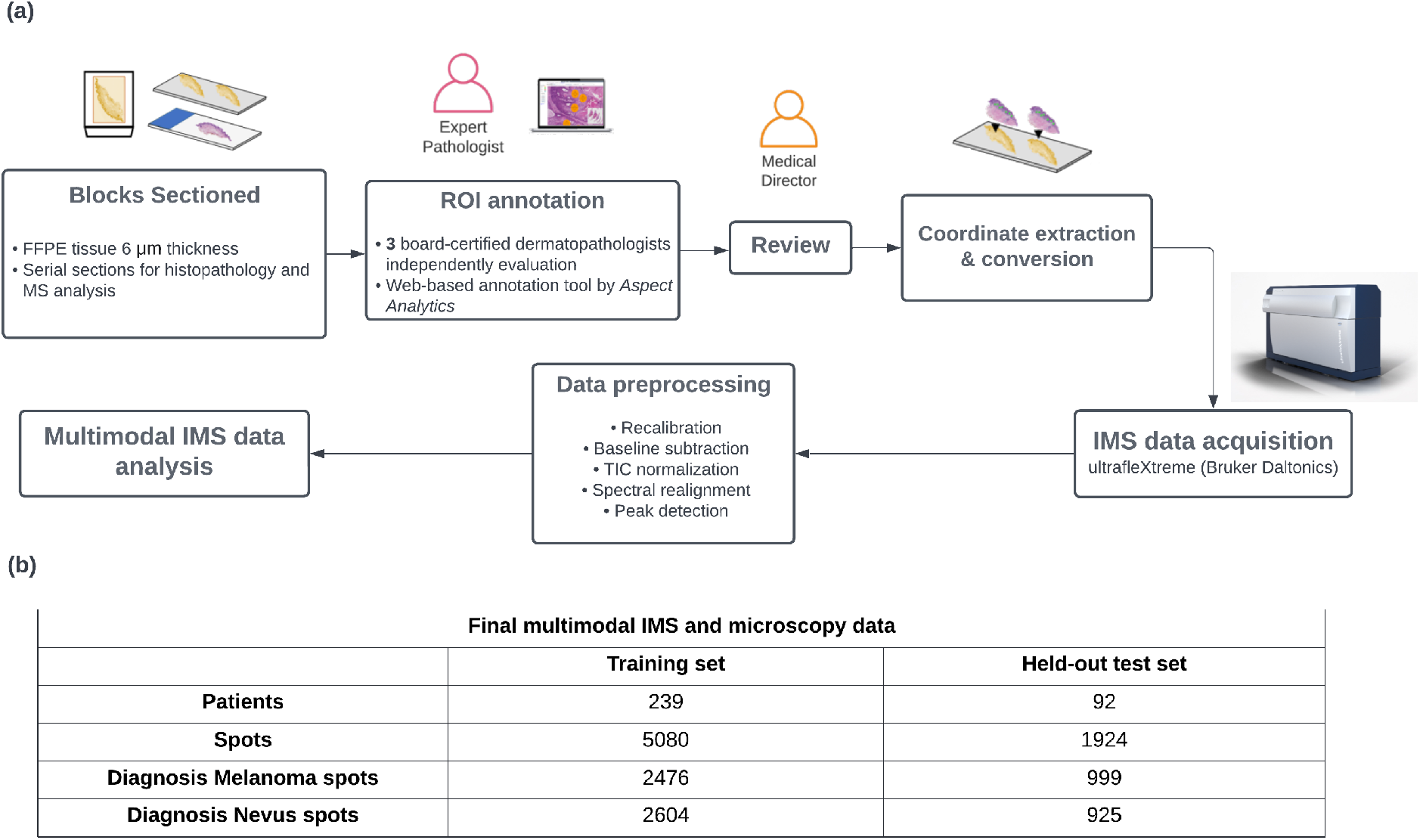
Overview of the histology-guided IMS sample preparation and processing workflow (a) and information about the final included number of samples and spots (b). The workflow (a) leads into multimodal data analysis and classification, incorporating both chemical information from MALDI IMS and morphological information from H&E microscopy.

Sections used for histopathological analysis were prepared and H&E stained according to standardized institutional practices and scanned at 20x magnification using a Leica SCN-400 digital slide scanner (Leica Biosystems, Buffalo Grove, IL, USA). The digitized sections were independently annotated by three board-certified dermatopathologists in a blinded fashion using web-based annotation tools: PIMS^23^ and Annotation studio (Aspect Analytics NV, Genk, Belgium). Each sample was provided to the pathologist with the patient demographic and with the goal of classifying the tissue as either melanoma or nevus.

Using the annotation software, the dermatopathologists annotated regions of interest from each sample/slide with a 50 µm diameter. While 6 different annotation labels were placed (melanoma *in situ*, invasive melanoma, junctional nevus, intradermal nevus, uninvolved epidermis and dermal stroma), this study focused only on building a binary classifier to distinguish the melanoma class from nevus class.

### MALDI IMS Sample Preparation and Data Acquisition

The sample preparation is the same as that described by Al-Rohil et al.. All sections for MALDI IMS were mounted on ITO slides (Delta Technologies, Loveland, CO). Briefly, sections were incubated in an oven at 55°C for 1 hour, deparaffinized (xylene x 2), rehydrated in ethanol (100% x 2, 95%, 70%), washed in ultrapure water (x2) and dried at 37°C. Antigen retrieval was conducted in EDTA buffer (pH 8.5) in an Instant Pot Electric Pressure cooker (IP-Duo60 V3) for 10 min on high pressure mode. For *in situ* tryptic digestion, sections were coated with 0.074 µg*/*mL mass spectrometry-grade trypsin (Sigma Aldrich) in a 100 mM ammonium bicarbonate buffer solution using a M5-Sprayer (HTX Technologies, Chapel Hill, NC), and incubated at 37 °C overnight. After digestion, the samples were coated in 5 mg/mL *α*-cyano-4-hydroxycinnamic acid matrix in 90% acetonitrile/deionized water and 0.2% TFA using the M5-Sprayer.

The dermatopathologist annotations were used to generate coordinates to guide the MALDI IMS acquisition from the same region/s on the unstained serial sections. MALDI IMS was performed on an ultrafleXtreme MALDI-TOF MS (Bruker Daltonics, Billerica, MA, USA) fitted with a SmartBeam Laser Nd:YAG laser following previously published parameters.^8^ Data was collected in positive reflector mode over the range of 700 to 3500 Daltons, at 500 laser shots per each profile spectrum for each region of interest.

### Data Processing and Sample Selection

A series of preprocessing methods were applied on the acquired IMS data, including resampling of the data to a common m/z axis, baseline subtraction, TIC normalization, spectral realignment and peak detection. ^8^

For the multi-modal image analysis, we only included tri-concordant samples (n = 331), meaning all dermatopathologists agreed with the diagnosis. Samples that were either diagnostically challenging, or did not meet the minimal mass spectral quality standards were excluded. Cases where only one modality is available (n=2) were also excluded from this study. Other than that, we used the same samples as the previously reported study by Al-Rohil et al., and can compare the multimodal results with the unimodal IMS classification reported therein.

The included samples contained 21 collected spots on average. As we focused on the binary classification task between melanoma and nevus, spots with annotations as melanoma (i.e., invasive melanoma and melanoma *in situ*) and nevus (i.e., junctional nevus and intradermal nevus) were included (n=7004). It must be noted that, in this study, we measured the classification performance only at the spots level and not at the aggregated patients/samples level. Our spot-level results can be directly compared to those reported in Al-Rohil et al..

The final multimodal IMS and microscopy dataset was randomly split into a training and a held-out test set at the patient level in order to prevent information leaks. Specifically, we designed our splits such that training and held-out test data spots originate from different patients, to ensure we are accurately assessing generalization performance on unseen slides, both during cross-validation fold generation (in the training data) as well as in determining the training vs held-out sets. The held-out test set in this study means that the data was never used during the whole model training part (during the parameter tuning, model selection). Figure 1(b) shows the information of the final included number of patients and the collected spots.

### Data Analysis

In this study, we built 3 different classification pipelines for melanoma diagnosis, namely the *unimodal MALDI IMS*, the *unimodal microscopy*, and the *multimodal pipeline*. As illustrated in Figure 2, the downstream classification part is the same across all compared pipelines but the inputs are different. For unimodal pipelines, only a single modality, either IMS data or microscopy, is utilized as the input for the downstream classification task, whereas in the multimodal setting, both modalities are used as the input for classification model training. Finally, these 3 pipelines are compared based on their classification performance.

**Figure 2:**
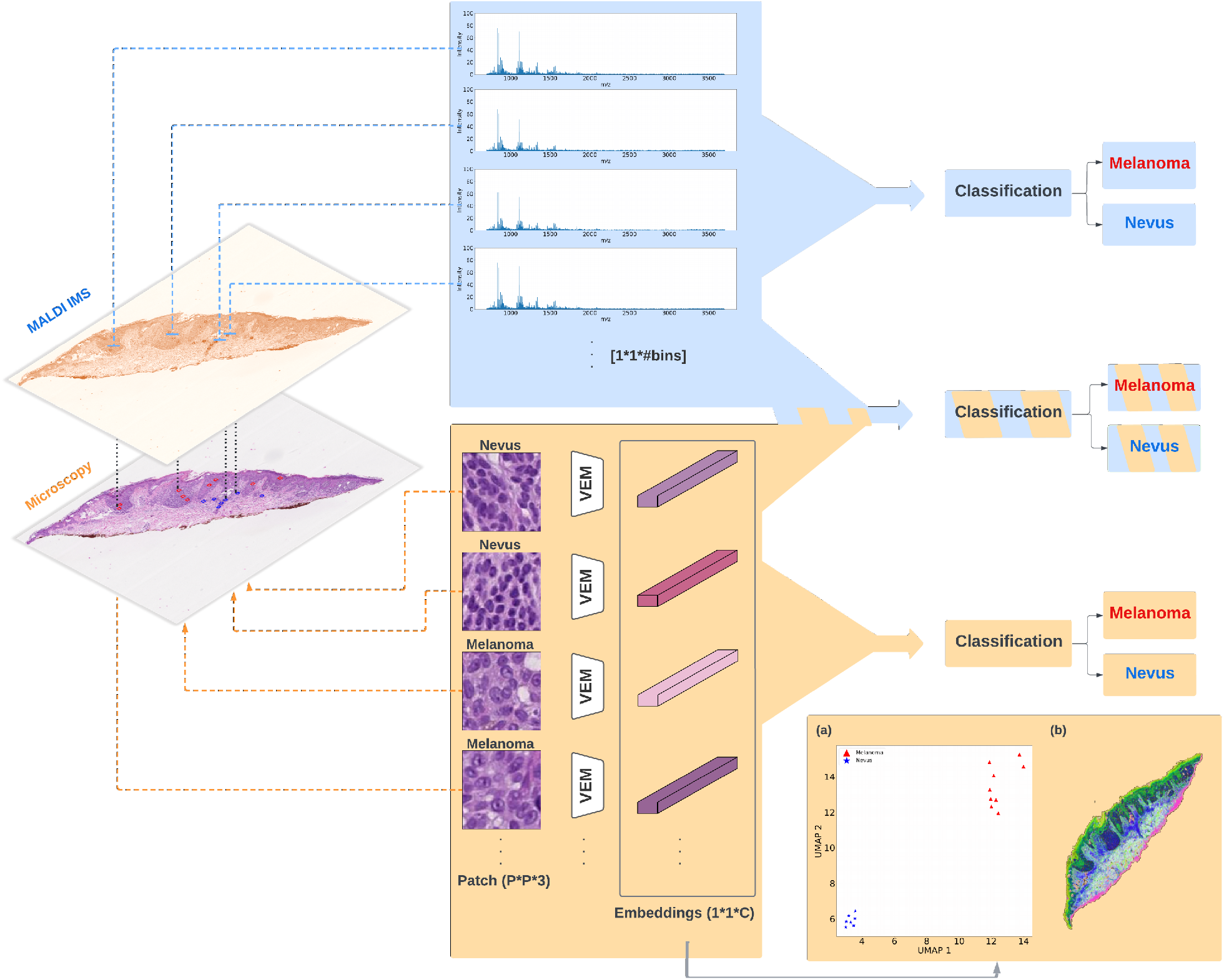
Data analysis workflow on an example tissue: **top blue panel** represents the unimodal IMS classification pipeline, where only IMS data are used for the downstream classification task; **bottom orange panel** shows the unimodal microscopy classification pipeline, where only morphology features are included for the melanoma diagnosis; In addition, intermediate embeddings from unimodal microscopy model were visualized via UMAP method: (a) 2-D embeddings of 16 patches (measured spots) from this example tissue with assigned colors based on pathologists’ annotations; (b) hyperspectral visualization of 3-D embeddings from all patches across the whole example tissue; **middle fused panel** is the multimodal strategy, where both IMS data and morphology features are used to distinguish melanoma from nevus. *#bins=5558, P=96, C=512, VEM: vector embedding morphology model*.

#### Unimodal IMS

Prior to feeding to the classification model, IMS data was preprocessed and peak picked. Each mass spectrum was generated from a single spot, which has 50 µm. After peak picking, a total number of 5558 peaks remained. Then we feed the preprocessed IMS data to the downstream classification model. This pipeline was reproduced from our previous study^8^ and compared with the multimodal pipeline in this study.

#### Unimodal Microscopy

We applied a pre-trained deep learning-based, self-supervised model to extract the features from microscopy data. ^20^ Each 3-color-channel histopathological image is transformed to a single dense vector and then fed to the downstream classification task. This pre-trained model was adapted originally from Chen et al., where the model learns the feature from data by maximizing the agreement between two stochastically augmented views of the same image via a contrastive loss function. Augmentation plays an essential role in contrastive learning. In the context of the histopathology dataset, some augmentation algorithms from the original paper^19^ may not be suitable here, as it was applied on natural-scene images like ImageNet.^24^ Thus, Ciga et al. conducted a series of experiments and some minor modifications on data augmentation. In the end, the augmentation algorithms included in the study are listed as following: randomly resized crops, 90 degrees rotations, horizontal and vertical flips, color jittering and Gaussian blurring.

Ciga et al. pre-trained their model on 57 multiple multi-organ histopathology datasets without any labels, as it is a self-supervised learning model. Those datasets contain various types of staining and resolutions, which helps the model learn better quality features. As reported in Ciga et al., the features learned from the model that was pre-trained on those histopathology datasets, are better than the features that were learned from networks pretrained on ImageNet. In total, more than 4 millions of patches (in pixel size of 224*224) are extracted from histopathology datasets and then fed to the model training.

In this study, we adapted this self-supervised model, pre-trained on a large number of histopathologic images, to the previously collected H&E stained histopathological images from our previous study.^8^ Each spot generates a single unique mass spectrum, and has a diameter of 50 µm. Thus, each patch in the histopathology dataset has a physical size of 48 µm*48 µm. The collection of histopathological patches consists of mainly of 20x resolution samples (96*96 pixel size), and a small portion of 40x resolution samples (192*192 pixel size). We applied some preprocessing steps on images following the suggestions in,^20^ including resizing, and normalization. Finally, each microscopy patch in the dataset is embedded into a vector representation via this pre-trained deep learning based self-supervised learning model, referred to as a *‘Vector Embedding Morphology’* (VEM) model in this paper. The resulting embeddings contain 512 dimensions. In order to visualize high-dimensional abstract features, we applied the method Uniform Manifold Approximation (UMAP)25 to project the data into lower dimensions. UMAP is a dimensionality reduction method, which seeks to search for a low-dimensional representation of the data that has the closest possible fuzzy topological representation of the original high-dimensional data. Previously, Smets et al. evaluated the performance of the UMAP method on IMS data and compared it with other popular dimensionality reduction algorithms like t-distributed stochastic neighbor embedding (t-SNE) and principal component analysis (PCA).

#### Multimodal IMS and Microscopy

In the multimodal setting, we represent the data for each spot with a single vector, which is created by concatenating the preprocessed IMS data for a spot together with the extracted morphological features from microscopy data for that same spot, with equal weights. In this way we achieve a fusing of the two modalities, which serves as an input for the classification algorithm.

#### Classification model

We applied Support Vector Machines (SVMs)^27,28^ to build the binary classifiers for each pipeline. As a popular supervised learning model, SVMs try to find a hyperplane as a decision boundary to separate data points with different classes, where this hyperplane has the maximal distance between data with different labels. To implement the SVMs algorithm, we used the Python package Scikit-learn^29^ with a linear kernel.

#### Model selection and evaluation

During the parameter tuning for SVMs models, we used a grid search strategy, where the model exhaustively generates candidates from a grid of parameter values that are predefined by the user. Furthermore, we leveraged *nested cross-validation* in order to estimate the generalization error underlying each model including the performance variability induced by the hyperparameter optimization process itself. Additionally, in order to further prevent information leaking, spots measured from the same patient/tissue were grouped in the same fold during the whole nest cross-validation process. In summary, we first split the total samples into a training set and a held-out test set on a patient level. Then we applied a grouped-nested-cross-validation strategy on the training set for model selection and evaluation. The held-out test set was not used during the whole training procedure. Thus the whole framework was designed carefully during the model selection and evaluation, so that we can fairly compare classification performance from unimodal and multimodal pipelines.

## Results

In this study, we built 3 different pipelines namely the unimodal microscopy, the unimodal IMS and the multimodal classification pipelines. Unimodal pipelines only use one modality i.e., either IMS or microscopy data for the downstream melanoma diagnosis, while multimodal strategy utilized both modalities. In order to compare those 3 pipelines, we conducted various measurements to assess their classification performance. In addition, we also applied a dimensionality reduction method UMAP to visualize the intermediate data in lower dimensions.

### UMAP visualization of intermediate data

In order to provide more intuition for these highly abstract features, we used UMAP to project and visualize the extracted morphology feature from the unimodal microscopy pipeline. As shown in Figure 2(a), after the feature extraction, each of those 16 patches (measured spots from the example tissue) is transformed into a single vector and shown in a 2-d scatter plot. Each dot in the scatter plot represents a vector representation of a microscopy patch and is assigned a red or blue color based on the pathology annotations either melanoma or nevus, respectively. Clearly, all melanoma patches are grouped together and far away from the cluster of nevus patches. Additionally, we collected all patches across the whole slide of this example tissue and used UMAP to project their 512-d transformed morphological embeddings into 3-d feature space. The resulting 3-d UMAP embeddings are visualized via translating each location of the patch to the RGB color scheme, which is referred to as hyperspectral visualization and shown in Figure 2(b). Regions where share similar colors represent their morphology are similar with each other. These results show the potential of the morphology features learned from *‘Vector Embedding Morphology’* (VEM) used in the unimodal microscopy pipeline, and provide more interpretation of these abstract embeddings. Importantly, this feature extraction process requires no training nor labels.

Furthermore, we performed UMAP on each input data from unimodal and multimodal pipelines. Three different inputs are collected and compared in this study, namely, (1) preprocessed IMS data (n_dimension = 5558), (2) morphological features extracted from microscopy data via a pre-trained self-supervised learning model (n_dimension = 512), (3) the combination of normalized (1) and (2) with equal weight (n_dimension = 5558+512 = 6070). UMAP was conducted to map those high-dimensional inputs to 2 dimensions in an unsupervised manner. Additionally, we also provide the labels for each data point in order to have a better intuition of the representations from those different input data.

As shown in Figure 3, each dot in the scatter plot represents a mass spectrum or a vector representation of a histopathological image or the combination of both, with its color showing the annotation provided by 3 dermatopathologists. Each column shows the UMAP embeddings of features from microscopy data, IMS and the combined data, respectively from left to right. The first and second rows represent the input data from the training dataset and the independent held-out test set respectively. We applied UMAP via Python implementation^30^ with cosine distance as metric for each type of input data.

**Figure 3:**
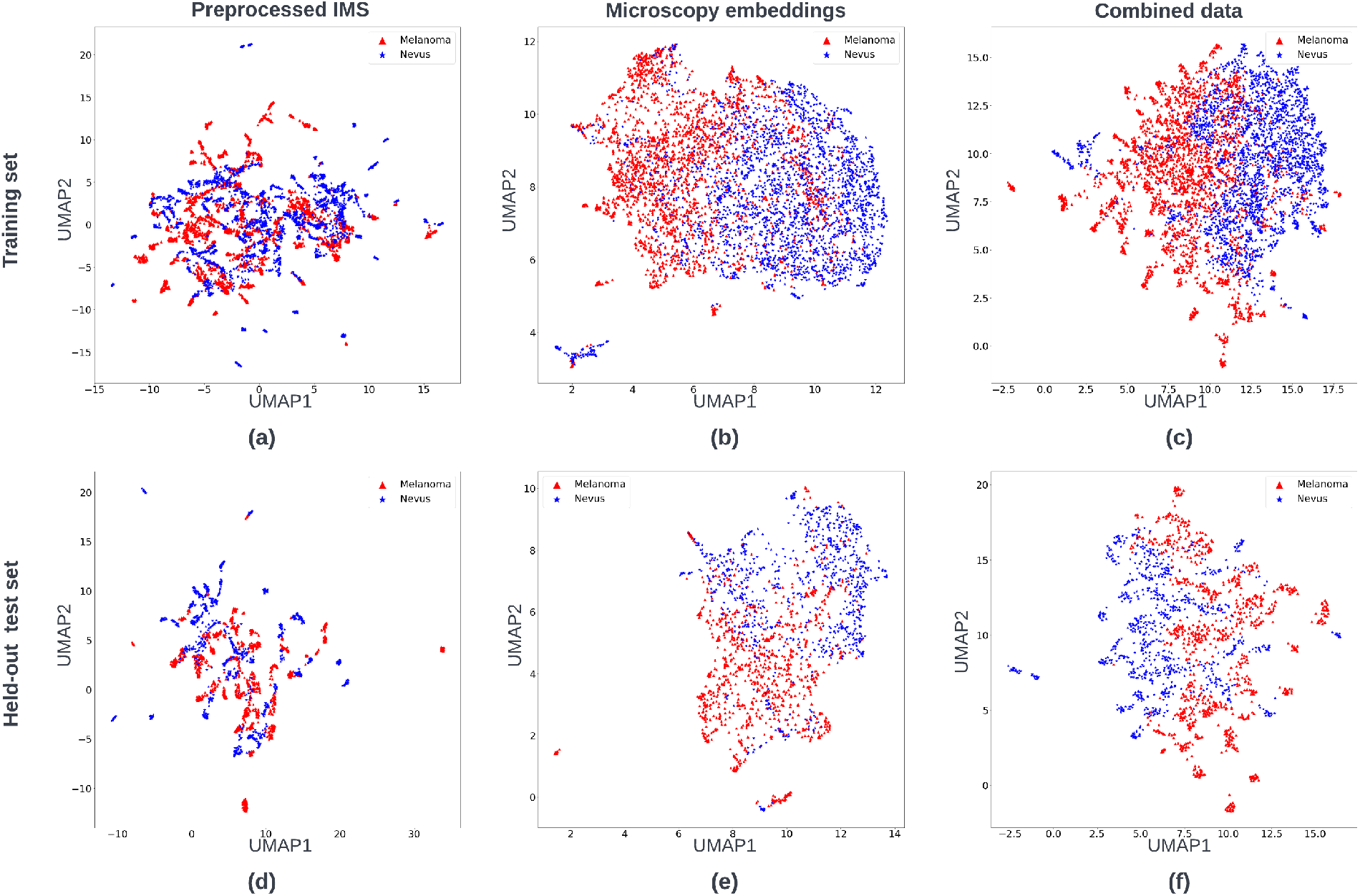
UMAP 2-D visualization of each input data across pipelines (unsupervised). In the first row (a) preprocessed IMS data, (b) extracted morphological features from microscopy data and (c) an equally weighted combination of (a) and (b) are from the training dataset. The second row shows the UMAP visualization from the held-out test dataset. The assigned color of each spot is decided by its diagnosis from 3 dermatopathologists.

### Classification performance

In this study we applied various classification performance measurements such as, Receiver operating characteristic - Area under the curve (ROC-AUC), F1, precision and recall scores. The range of those metrics is typically from 0 to 1, where 1 represents the perfect classification performance. ROC curves are widely used to measure performance in binary classification tasks, as it plots the true positive rates (TPR) and the false positive rate (FPR) with different thresholds. By computing the area under the ROC curve, the performance across the full range of classification thresholds can be summarized in a single number.

As mentioned earlier, we carefully performed the *grouped-nest-cross-validation* strategy throughout the modeling process in order to avoid any possible information leaking. We applied this strategy on the training dataset and fit the optimized classification model for each pipeline on an independent held-out test set. The results are shown in Figure 4.

**Figure 4:**
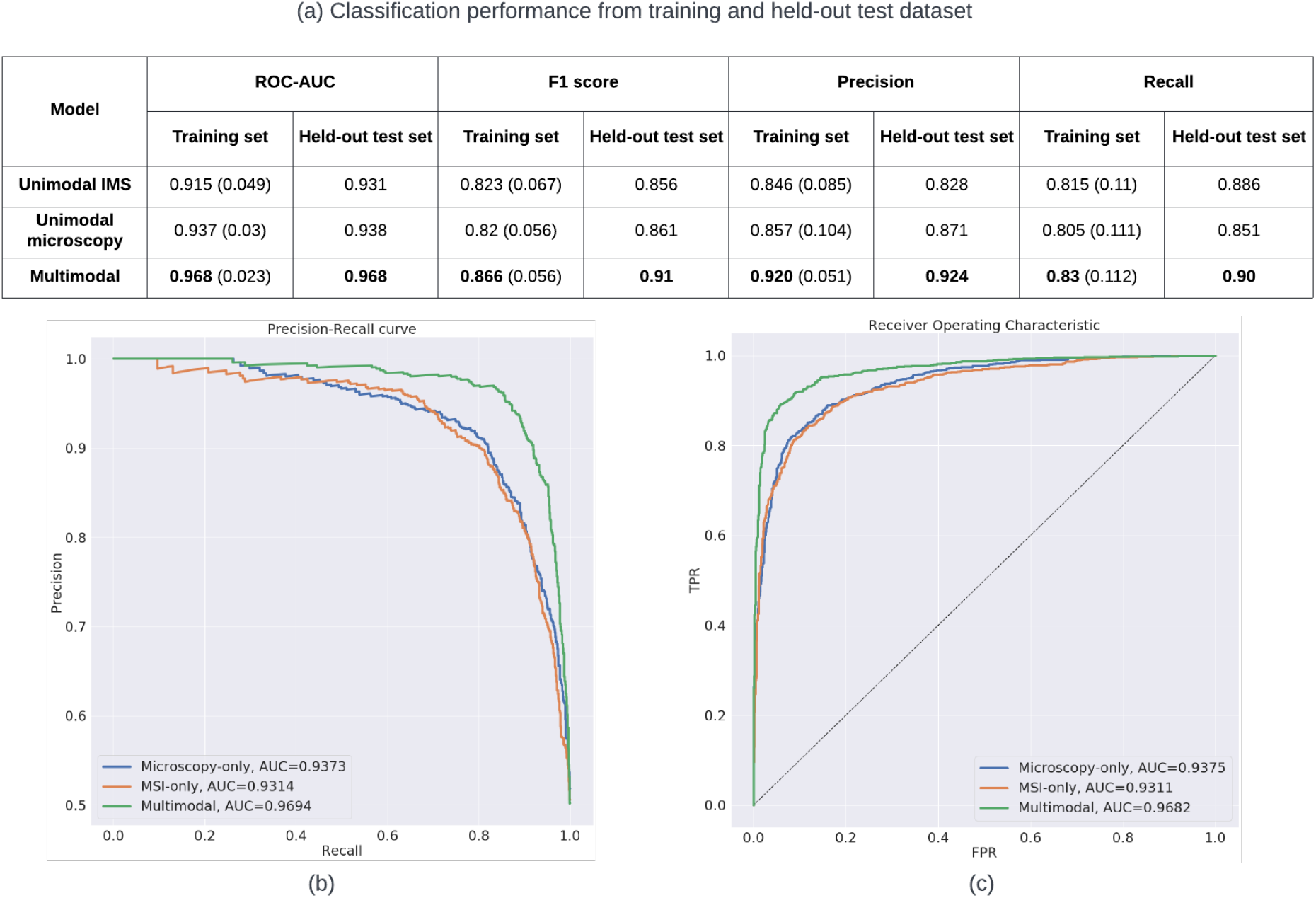
Classification results on training and held-out test sets. All results are based on **spot-level**. (a) shows the classification performance of each pipeline on training and held-out test dataset. The results from the training set represent the mean value of the performance (with their standard deviation) after the nested-cross-validation process. (b)and(c) show the precision-recall and ROC cures plots of each pipeline on the **held-out test set**.

We reproduced the unimodal MALDI IMS classification results from our previous study, ^8^ and obtained comparable ROC-AUC results (0.910 vs. 0.915 on training set, 0.933 vs. 0.938 on the held-out test set, for reported spots-level results from Al-Rohil et al. and our reproduced results, respectively). Interestingly, the unimodal microscopy pipeline achieves higher mean ROC-AUC scores than the unimodal IMS (0.937 vs. 0.915) on the training dataset, while achieving comparable ROC-AUC results on the held-out test set. The result shows the advantage of using a pre-trained deep learning model for extracting important morphological features from microscopy data. However, it must be noted that the annotations used to train and evaluate the classification model are fully based on microscopy data by dermatopathologists.

By combining information originating from the two available modalities in a single classifier, i.e. the multimodal pipeline, we obtain the best classification performance, (ROC-AUC as 0.968 on training and held-out test set). This demonstrates the synergy in combining observations in the biochemical domain, obtained through IMS, with those in the morphology domain, which have been traditionally used in melanoma diagnosis.

The unimodal IMS and unimodal microscopy pipelines show comparable F1 scores across both training and held-out test sets. Unimodal IMS outperforms unimodal microscopy on both training and held-out test sets based on the recall scores (0.815 vs. 0.805, 0.886 vs. 0.851). On the other hand, unimodal microscopy achieves higher precision scores than unimodal IMS on training and held-out test sets. These slight variations in terms of which approach is best are attributable to differences in classification thresholds, thus emphasizing the importance of using an overall metric like ROC-AUC that accounts for the classifiers’ full range.

We also computed the standard deviation for each score based on the training data, and the multimodal pipeline achieves the lowest values almost across all metrics. For example, standard deviations from ROC-AUC: 0.023 (multimodal) vs 0.03 (unimodal microscopy) vs 0.049 (unimodal IMS). This indicates higher stability of the multimodal model, which additionally reinforces the value of combining both modalities.

Additionally, we plotted the Precision-Recall (PR) curve and ROC curves from the independent held-out test set in Figure 4(b) and (c) respectively. PR curves illustrate the trade-off between precision score and recall score with different thresholds. Similarly to ROC-AUC, higher area under the PR curve (PR-AUC) represents higher recall and precision across all classification thresholds. PR and ROC curves are typically used to assess the binary classification performance. As shown in Figure 4(b)(c), the multimodal pipeline achieves the highest PR-AUC and ROC-AUC scores (0.969, 0.968).

Furthermore, we encountered some interesting misclassified cases on the held-out test dataset. As shown in Figure 5, the multimodal strategy shows its power in accurate classification of melanoma and nevus, even when both unimodal strategies fail to yield correct predictions. This highlights that our multimodal classification model is greater than the sum of its (unimodal) parts.

**Figure 5:**
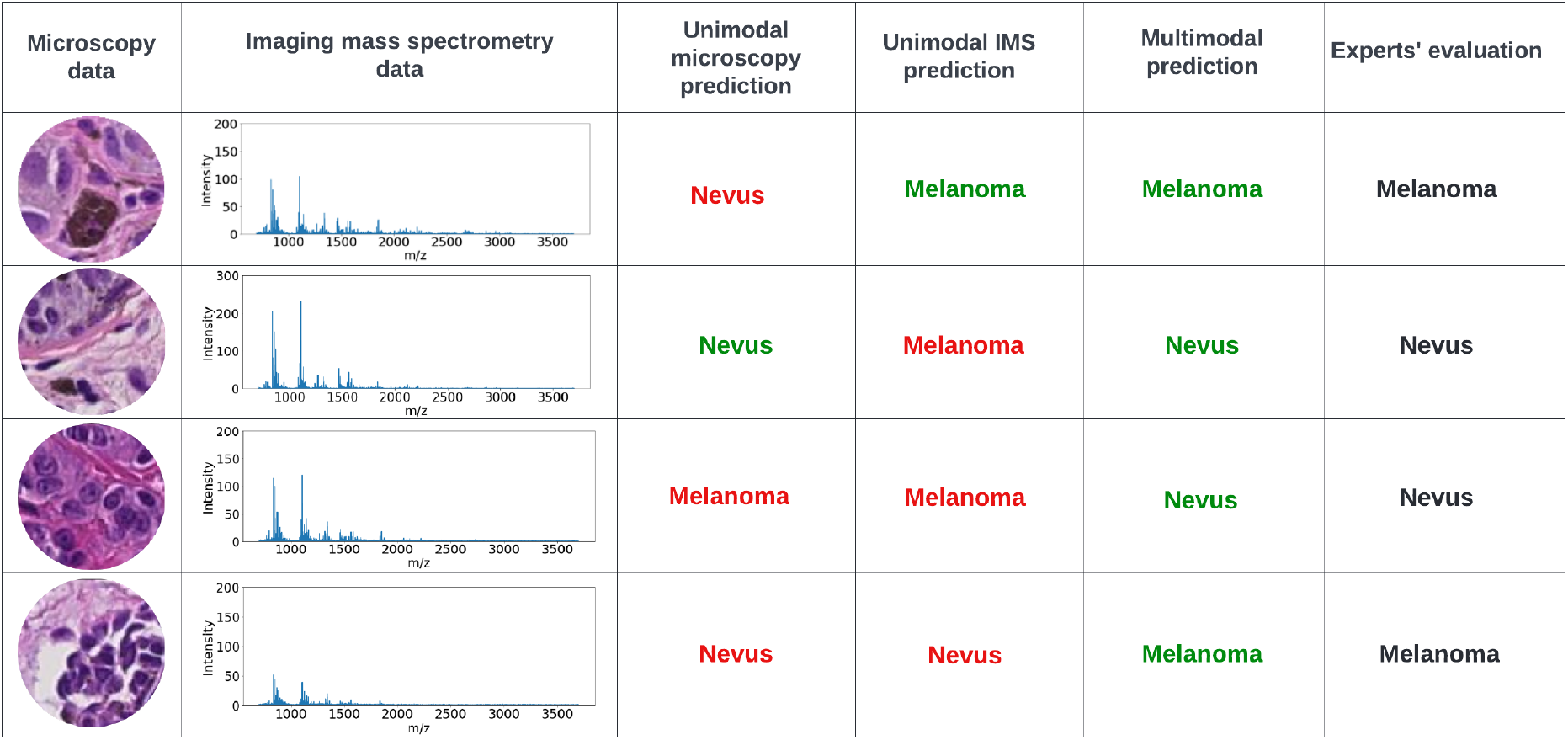
Misclassified cases by unimodal algorithms from the held-out test set. Red and green text indicate model results that are incongruent and congruent with expert pathology assessment, respectively.

## Discussion

This study extends and improves the previous melanoma diagnosis study by Al-Rohil et al. by incorporating the morphological features from microscopy data in the classification model along with MALDI IMS data. The spot-level ROC-AUC results from the classification model have been largely improved from previous study strategy namely unimodal MALDI IMS (0.915 vs. 0.968, training data), as well as the other scores such as F1 score, precision and recall. Additionally, the classification model training became more stable and robust with the help of multimodal strategy, evidenced by lower variability in model performance.

At present, most IMS classification studies largely focus on only using histopathology data to make annotations that guide the IMS data collection or restrict which spectra are used in order to build more accurate classifiers.^31,32^ Secondly, multimodal MALDI IMS analysis requires careful data acquisition, dedicated registration before the downstream data analysis, which is not always trivial. Last but not the least, learning high-quality morphological features from microscopy data could require a large amount of manpower for dedicated annotations and also introduce computational challenges.

To our knowledge, there is only one study^17^ utilizing the information from histopathology data, via computer-aided morphometric-based image analysis, in combination with MALDI IMS to improve renal carcinoma subtyping. In comparison with their work, instead of using morphometric-based algorithms, we applied a deep learning-based self-surprised model (pretrained on a large histopathology dataset) to extract the high-quality morphological features from microscopy data in a fully unsupervised manner. This model is highly efficient as there is no training required during the feature extraction. However, the extracted features are highly abstract, which is a common challenge in the deep learning field. Thus the resulting features from microscopy data are not as interpretable as the ones learned from the morphometric-based method as used by Prade et al.. To improve the interpretability of the learned features, we applied the dimensionality reduction method like UMAP on those high-dimensional features (512) and projected them into 2 & 3-d feature space.

Finally, we built a linear classifier on the top of IMS features and/or morphological features to distinguish melanoma from nevus. The classification results show that a multimodal strategy, with a relatively simple equal weighted combination method, can outperform other unimodal workflows in both prediction accuracy and robustness. Microscopy images are acquired in tandem in a majority of IMS experiments, and as such this methodology provides excellent opportunities to improve classification and downstream machine learning at very little additional expense. Due to the use of pre-trained networks, which do not require additional training, this additional feature extraction step from microscopy images is near ‘plug-and-play’, and could be readily incorporated with IMS experiments on other types of tissues in the future.

## Conclusion

With the increased prevalence of multimodal studies, especially those integrating traditional histological or cellular stains with molecular analyses, it is important to develop computational approaches that effectively combine the data to go beyond qualitative comparisons. A critical area of such combinations is clinical assays where the increase in information from modality combinations has the potential to improve health outcomes. In this work, we demonstrate the potential of a multimodal strategy on a relatively large patient cohort (n=331) which combines morphological features extracted from standard clinical microscopy data and MALDI IMS molecular data the melanoma diagnosis. In future work, this strategy can be further validated on other types of cancer tissues, more advanced fusion strategies could be developed, and the approach could be applied to other combinations of morphological optical images and molecular data.

## Data Availability

The data on which this study is based cannot be disclosed.

## Acknowledgement

The authors thank Ahmed Alomari, Rami Al-Rohil, Jason Robbins for for providing pathological analysis, and Thomas Moerman for software development. This work was supported by Small Business Innovative Research Grant No. R44CA228897-02, provided by the National Cancer Institute of the National Institutes of Health. Wanqiu Zhang was supported by a Baekeland PhD grant from Flanders Innovation and Entrepreneurship (VLAIO). KU Leuven: Research Fund (projects C16/15/059, C3/19/053, C24/18/022, C3/20/117, C3I-21-00316), Industrial Research Fund (Fellowships 13-0260, IOFm/16/004) and several Leuven Research and Development bilateral industrial projects; Flemish Government Agencies: FWO: EOS Project no G0F6718N (SeLMA), SBO project S005319N, Infrastructure project I013218N, TBM Project T001919N; PhD Grant (SB/1SA1319N), EWI: the Flanders AI Research Program, VLAIO: CSBO (HBC.2021.0076) Baekeland PhD (HBC.20192204), European Commission: European Research Council under the European Union’s Horizon 2020 research and innovation programme (ERC Adv. Grant grant agreement No 885682); Other funding: Foundation ‘Kom op tegen Kanker’, CM (Christelijke Mutualiteit).

## References

(1) https://www.cancer.org/cancer/melanoma-skin-cancer/about/key-statistics.html.

(2) Leiter, U.; Keim, U.; Garbe, C. Epidemiology of skin cancer: update 2019. Sunlight, Vitamin D and Skin Cancer 2020, 123–139.

(3) Gerami, P.; Jewell, S. S.; Morrison, L. E.; Blondin, B.; Schulz, J.; Ruffalo, T.; Matushek IV, P.; Legator, M.; Jacobson, K.; Dalton, S. R., et al. Fluorescence in situ hybridization (FISH) as an ancillary diagnostic tool in the diagnosis of melanoma. The American journal of surgical pathology 2009, 33, 1146–1156.

(4) Veenhuizen, K. C.; De Wit, P. E.; Mooi, W. J.; Scheffer, E.; Verbeek, A. L.; Ruiter, D. J. Quality assessment by expert opinion in melanoma pathology: experience of the pathology panel of the Dutch Melanoma Working Party. The Journal of Pathology: A Journal of the Pathological Society of Great Britain and Ireland 1997, 182, 266–272.

(5) Zembowicz, A.; Scolyer, R. A. Nevus/melanocytoma/melanoma: an emerging paradigm for classification of melanocytic neoplasms? Archives of pathology & laboratory medicine 2011, 135, 300–306.

(6) Porta Siegel, T.; Hamm, G.; Bunch, J.; Cappell, J.; Fletcher, J. S.; Schwamborn, K. Mass spectrometry imaging and integration with other imaging modalities for greater molecular understanding of biological tissues. Molecular Imaging and Biology 2018, 20, 888–901.

(7) Casadonte, R.; Kriegsmann, M.; Kriegsmann, K.; Hauk, I.; Meliß, R. R.; Müller, C. S.; Kriegsmann, J. Imaging mass spectrometry-based proteomic analysis to differentiate melanocytic nevi and malignant melanoma. Cancers 2021, 13, 3197.

(8) Al-Rohil, R. N.; Moore, J. L.; Patterson, N. H.; Nicholson, S.; Verbeeck, N.; Claesen, M.; Muhammad, J. Z.; Caprioli, R. M.; Norris, J. L.; Kantrow, S., et al. Diagnosis of melanoma by imaging mass spectrometry: Development and validation of a melanoma prediction model. Journal of Cutaneous Pathology 2021, 48, 1455–1462.

(9) Lazova, R.; Seeley, E. H. Proteomic mass spectrometry imaging for skin cancer diagnosis. Dermatologic Clinics 2017, 35, 513–519.

(10) Taverna, D.; Nanney, L. B.; Pollins, A. C.; Sindona, G.; Caprioli, R. Spatial mapping by imaging mass spectrometry offers advancements for rapid definition of human skin proteomic signatures. Experimental dermatology 2011, 20, 642–647.

(11) de Macedo, C. S.; Anderson, D. M.; Schey, K. L. MALDI (matrix assisted laser desorption ionization) imaging mass spectrometry (IMS) of skin: aspects of sample preparation. Talanta 2017, 174, 325–335.

(12) Guran, R.; Vanickova, L.; Horak, V.; Krizkova, S.; Michalek, P.; Heger, Z.; Zitka, O.; Adam, V. MALDI MSI of MeLiM melanoma: Searching for differences in protein profiles. PLoS One 2017, 12, e0189305.

(13) Lazova, R.; Smoot, K.; Anderson, H.; Powell, M. J.; Rosenberg, A. S.; Rongioletti, F.; Pilloni, L.; D’Hallewin, S.; Gueorguieva, R.; Tantcheva-Poór, I., et al. Histopathology-guided mass spectrometry differentiates benign nevi from malignant melanoma. Journal of Cutaneous Pathology 2020, 47, 226–240.

(14) Alomari, A. K.; Glusac, E. J.; Choi, J.; Hui, P.; Seeley, E. H.; Caprioli, R. M.; Watsky, K. L.; Urban, J.; Lazova, R. Congenital nevi versus metastatic melanoma in a newborn to a mother with malignant melanoma–diagnosis supported by sex chromosome analysis and imaging mass spectrometry. Journal of Cutaneous Pathology 2015, 42, 757–764.

(15) Taverna, D.; Boraldi, F.; De Santis, G.; Caprioli, R. M.; Quaglino, D. Histology-directed and imaging mass spectrometry: An emerging technology in ectopic calcification. Bone 2015, 74, 83–94.

(16) Tuck, M.; Grélard, F.; Blanc, L.; Desbenoit, N. MALDI-MSI Towards Multimodal Imaging: Challenges and Perspectives. Frontiers in Chemistry 2022, 10.

(17) Prade, V. M.; Sun, N.; Shen, J.; Feuchtinger, A.; Kunzke, T.; Buck, A.; Schraml, P.; Moch, H.; Schwamborn, K.; Autenrieth, M., et al. The synergism of spatial metabolomics and morphometry improves machine learning-based renal tumour subtype classification. Clinical and translational medicine 2022, 12.

(18) Erlmeier, F.; Feuchtinger, A.; Borgmann, D.; Rudelius, M.; Autenrieth, M.; Walch, A. K.; Weirich, G. Supremacy of modern morphometry in typing renal oncocytoma and malignant look-alikes. Histochemistry and cell biology 2015, 144, 147–156.

(19) Chen, T.; Kornblith, S.; Norouzi, M.; Hinton, G. A simple framework for contrastive learning of visual representations. International conference on machine learning. 2020; pp 1597–1607.

(20) Ciga, O.; Xu, T.; Martel, A. L. Self supervised contrastive learning for digital histopathology. Machine Learning with Applications 2022, 7, 100198.

(21) Verbeeck, N.; Caprioli, R. M.; Van de Plas, R. Unsupervised machine learning for exploratory data analysis in imaging mass spectrometry. Mass spectrometry reviews 2020, 39, 245–291.

(22) Liu, X.; Zhang, F.; Hou, Z.; Mian, L.; Wang, Z.; Zhang, J.; Tang, J. Self-supervised learning: Generative or contrastive. IEEE Transactions on Knowledge and Data Engineering 2021,

(23) Norris, J. L.; Tsui, T.; Gutierrez, D. B.; Caprioli, R. M. Pathology interface for the molecular analysis of tissue by mass spectrometry. Journal of Pathology Informatics 2016, 7, 13.

(24) Deng, J.; Dong, W.; Socher, R.; Li, L.-J.; Li, K.; Fei-Fei, L. Imagenet: A large-scale hierarchical image database. 2009 IEEE conference on computer vision and pattern recognition. 2009; pp 248–255.

(25) McInnes, L.; Healy, J.; Melville, J. Umap: Uniform manifold approximation and projection for dimension reduction. arXiv preprint 1802.03426 2018,

(26) Smets, T.; Verbeeck, N.; Claesen, M.; Asperger, A.; Griffioen, G.; Tousseyn, T.; Waelput, W.; Waelkens, E.; De Moor, B. Evaluation of distance metrics and spatial autocorrelation in uniform manifold approximation and projection applied to mass spectrometry imaging data. Analytical chemistry 2019, 91, 5706–5714.

(27) Cortes, C.; Vapnik, V. Support-vector networks. Machine learning 1995, 20, 273–297.

(28) Luts, J.; Ojeda, F.; Van de Plas, R.; De Moor, B.; Van Huffel, S.; Suykens, J. A. A tutorial on support vector machine-based methods for classification problems in chemometrics. Analytica chimica acta 2010, 665, 129–145.

(29) Pedregosa, F.; Varoquaux, G.; Gramfort, A.; Michel, V.; Thirion, B.; Grisel, O.; Blondel, M.; Prettenhofer, P.; Weiss, R.; Dubourg, V., et al. Scikit-learn: Machine learning in Python. the Journal of machine Learning research 2011, 12, 2825–2830.

(30) McInnes, L.; Healy, J.; Saul, N.; Grossberger, L. UMAP: Uniform Manifold Approximation and Projection. The Journal of Open Source Software 2018, 3, 861.

(31) Deininger, S.-O.; Bollwein, C.; Casadonte, R.; Wandernoth, P.; Gonçalves, J. P. L.; Kriegsmann, K.; Kriegsmann, M.; Boskamp, T.; Kriegsmann, J.; Weichert, W., et al. Multicenter Evaluation of Tissue Classification by Matrix-Assisted Laser Desorption/Ionization Mass Spectrometry Imaging. Analytical Chemistry 2022,

(32) Gonçalves, J. P. L.; Bollwein, C.; Schlitter, A. M.; Martin, B.; Märkl, B.; Utpatel, K.; Weichert, W.; Schwamborn, K. The impact of histological annotations for accurate tissue classification using mass spectrometry imaging. Metabolites 2021, 11, 752.

